# Genetic drivers of progression in Alzheimer’s disease are distinct from disease risk

**DOI:** 10.1101/2025.11.17.25340247

**Authors:** Celeste E Cohen, Shane Fernandez, Umran Yaman, Ahmad R Ehyaei, Tenielle Porter, Eleanor O’Brien, Australian Imaging Biomarkers and Lifestyle Study, Alzheimer’s Disease Neuroimaging Initiative, Paul Maruff, John A. Hardy, Simon M. Laws, Dervis Salih, Maryam Shoai

**Author notes:** Contributed equally (Celeste E Cohen, Shane Fernandez). Data used in the preparation of this article were obtained from the Australian Imaging, Biomarker and Lifestyle (AIBL) Study database [https://aibl.org.au/collaboration]. As such, the investigators within AIBL, unless listed, contributed to the design and implementation of AIBL and/or provided data but did not participate in the analysis or writing of this report. A complete listing of AIBL investigators can be found at: [https://aibl.org.au/about/our-researchers/]. Data used in preparation of this article were obtained from the Alzheimer’s Disease Neuroimaging Initiative (ADNI) database (adni.loni.usc.edu). As such, the investigators within the ADNI contributed to the design and implementation of ADNI and/or provided data but did not participate in the analysis or writing of this report. A complete listing of ADNI investigators can be found at: [http://adni.loni.usc.edu/wpcontent/uploads/how].

## Abstract

**Background:** Recent trials in Alzheimer’s disease (AD) demonstrate encouraging outcomes. These trials target risk mechanisms identified through genetic analysis whilst directly aiming to reduce progression rates. Evidence from other neurodegenerative diseases suggests the genetics of progression is distinct from risk of disease. To expand these initial successes and improve clinical outcomes further we need to understand genetics of progression of disease. These can be deduced through rigorous analysis of meticulously phenotyped longitudinal cohorts. In this study we first looked at known genetic drivers of risk, namely polygenic risk scores for AD and *APOE*–ε*4*, to assess their role in progression. This was then extended to a genome wide association analysis to identify the role of other genetic variants in progression of AD.

**Methods:** A total of 387 individuals with, genetic data, amyloid positivity and in active decline (ADNI (n=222) and AIBL(n=165)) were used to perform generalised mixed effects linear model genome wide association studies of longitudinal cognitive decline as measured by mini mental state examination. The resulting summary statistics were subjected z, and colocalization analyses.

**Results:** Established AD risk factors, including *APOE*–ε*4* dosage and polygenic risk scores, were not associated with disease progression amyloid positive individuals who are actively declining. A mixed effects GWAS meta-analysis revealed one genome-wide significant locus on chromosome 22 (rs78369883) and 25 nominally significant loci linked with AD progression. Functional annotation, finemapping, and colocalization analyses implicated genes primarily involved in immune response, neurodegeneration (including tau pathology), brain resilience, and neurogenesis. These progression-related genes were significantly enriched in neuronal-interferon-microglial signalling pathways and normal homeostatic processes of neuronal networks, with specific enrichment in dopaminergic and inhibitory neuronal populations.

**Conclusion:** These findings enhance our understanding of the biological underpinnings of AD progression, opening new avenues for therapeutic intervention.

## 1. Introduction

Genetic studies have been central to elucidation of the pathogenesis of Alzheimer’s disease (AD), including analyses of familial early-onset cases, genome-wide association studies (GWAS), and transgenic models. These studies have identified genes associated with the development of AD generally, and those related directly to the formation of the canonical neuropathological hallmarks of AD: plaques composed of misfolded amyloid-β (Aβ) peptides and neurofibrillary tangles (NFTs) of hyperphosphorylated tau proteins [1]. Identification and modification of these pathways has become the objective of pharmacotherapies developed to prevent or slow the progression of AD. Of these, monoclonal antibodies which influence the rate of accumulation of Aβ plaques, aducanumab, lecanemab, and donanemab have been approved for treatment of AD [2–4]. These drugs produce modest clinical benefit (∼30% slowing of progression) in adults with early symptomatic AD, although accumulating evidence suggests this benefit can increase with time on treatment, especially in early disease. Monoclonal antibodies are also associated with increased risk of adverse events, especially, amyloid-related imaging abnormalities (ARIA) [4–6]. These outcome characteristics indicate the need for greater understanding of relationships between drugs designed to influence amyloid, including their dose, dosing schedules, and the extent of amyloid accumulation in the target populations [7].

Disease pathways other than those that target Aβ are also under investigation. Tau pathology is a strong candidate as its accumulation correlates well with clinical disease severity and progression [8, 9]. Clinical trials of drugs designed to remove tau have thus far failed to meet endpoints or are still underway [10]. This leaves a critical gap in identifying viable therapeutic targets for disease progression.

It is important to acknowledge that clinical trials have primarily leveraged an understanding of disease pathogenesis, exemplified by the involvement of Aβ and Tau, to inform the design of therapeutic targets. These trials rigorously evaluate the efficacy of these targeted interventions in cohorts of patients *after disease onset*, with the objective of attenuating disease progression. Implicit in this approach, is the underlying assumption that the biological mechanisms precipitating disease onset are the same as those dictating the rate of disease progression. However, in the context of neurodegenerative disorders such as Parkinson’s disease [11] and Progressive Supranuclear Palsy [12], evidence from GWAS has demonstrated a divergence: the genetic variants associated with disease susceptibility (risk GWAS) are distinct from those that drive progression of disease as seen in progression GWAS.

It is probable that therapeutic targets derived from biology of disease progression can add major insight in slowing or halting of disease.

To identify novel therapeutic targets, it is essential to distinguish causal drivers of pathogenesis from progression. While pathways involving mitochondrial dysfunction or inflammation have been linked to rapid progression, these proteomic and physiological associations may be downstream effects of true drivers of progression [13–15]. Genetic studies are positioned uniquely to identify other causal drivers of disease. While genetic pathways other than those related to amyloid and tau have been identified, validation and exploitation of these relationships has been hampered by methodological limitations, such as phenotype impurity, assessing known risk genes only, and not accounting for interpatient heterogeneity. Candidate gene analyses such as *MAPT* and *PPP3R1* are informative but very limited [16]. Lastly, genome wide analyses have often been confounded by the absence of disease confirmatory biomarkers such as amyloid, which is critical given the high rate of clinical misdiagnosis[17–19]. Furthermore, absence of long-term prospective data with multiple follow up points often gives a very biased snapshot of disease.

To overcome these limitations, a rigorous methodological framework is required. Progression in AD is typically measured by cognitive decline, and the Mini-Mental State Examination (MMSE) has demonstrated greater response to change than other scales used in prior GWAS [20]. For modelling this longitudinal data, linear mixed-effects models are methodologically superior, as they best account for between and inter-patient heterogeneity [21].

In this study we attempt to uncouple the genetics of AD risk and progression. Using two amyloid-positive cohorts, we first examined whether rates of progression were associated with established AD risk factors; *APOE*–ε*4* dosage and polygenic risk scores. We then conducted a comprehensive GWAS meta-analysis of cognitive decline, measured by MMSE with linear mixed-effects models to identify genetic factors that may influence rate of progression, and thus move towards identification of therapeutic targets that may provide alternative mechanisms for treatment of AD.

## 2. Methods

### 2.1. Cohorts

#### The Alzheimer’s Disease Neuroimaging Initiative (ADNI)

ADNI comprises genomic, imaging, biospecimen, and longitudinal data from mainly North American individuals to study AD progression [22]. Clinical (15,765 visits of 2,379 patients in ADNIMERGE) and genetic (809 patients across ADNI and 757 from ADNI1) data were downloaded from the Image and Data Archive at the Laboratory of Neuro Imaging (IDA LONI) [23] for three different ADNI studies: ADNI 1, GO and 2.

#### The Australian Imaging, Biomarker and Lifestyle (AIBL) study of aging

The flagship Study of Ageing collects genomic, imaging, biospecimen and longitudinal clinical data from centres in Perth and Melbourne, Australia to study AD onset and progression [24, 25] in 2359 older adults with visits scheduled every 18 months.

#### Ethical compliance

All human procedures complied with the ethical standards of the responsible committees on human experimentation (institutional and national) and with the Helsinki Declaration of 1975, as revised. Full ethical approval and informed consent were obtained by the respective ADNI and AIBL institutions for all participants and subsequent public release of anonymized data.

As a secondary analysis of de-identified, publicly accessible data, the study was deemed exempt from additional institutional review board review. All participants provided written informed consent to donate their biospecimens and clinical data for use in future research studies

### 2.2. Genomic quality control and imputation

Both datasets were aligned to the GRCh37/hg19 reference genome.

Quality control (QC) was performed on ADNI whole genome sequenced data, ADNI genotyped data and AIBL genotyping datasets using PLINK 1.9 and 2.0 [26, 27]. Data QC was performed per-individual then per-locus, with the most stringent criteria possible to maintain confidence whilst maximising sample numbers. Given that ADNI and AIBL subjects are overwhelmingly Caucasian, only these were retained to avoid population stratification due to differences in ancestry. Ancestry was deduced using principal component analysis against HapMap [28] in ADNI, and 1000 genomes in AIBL.

Briefly per-individual QC comprised checks for excessive heterozygosity, genotype missingness, reported and genetic sex discordance, relatedness, and ancestry.

SNP QC was performed to remove SNPs with deviations from Hardy Weinberg Equilibrium at *p* ≤1 ×10^-5^, missing genotyping rate above 1%, and minor allele frequency below 1%.

Imputation of ADNI data was done via the Sanger Imputation Service [29] for phasing (EAGLE2) and imputation (PBWT), using UK 10K [30] and 1000 Genomes Phase 3 haplotype data [31]. SNPs with imputation score less than 0.7 were removed after imputation. AIBL data was uploaded to the TOPMed Imputation Server Pipeline for phasing (Eagle v2.4) and imputation (Minimac4). Imputed variants with R^2^ < 0.3 were removed by TOPMed. Both imputed datasets were then subjected to post imputation QC in line with the genetic and individual QC steps carried out prior to imputation as detailed in Figure 1.

**Figure 1.**
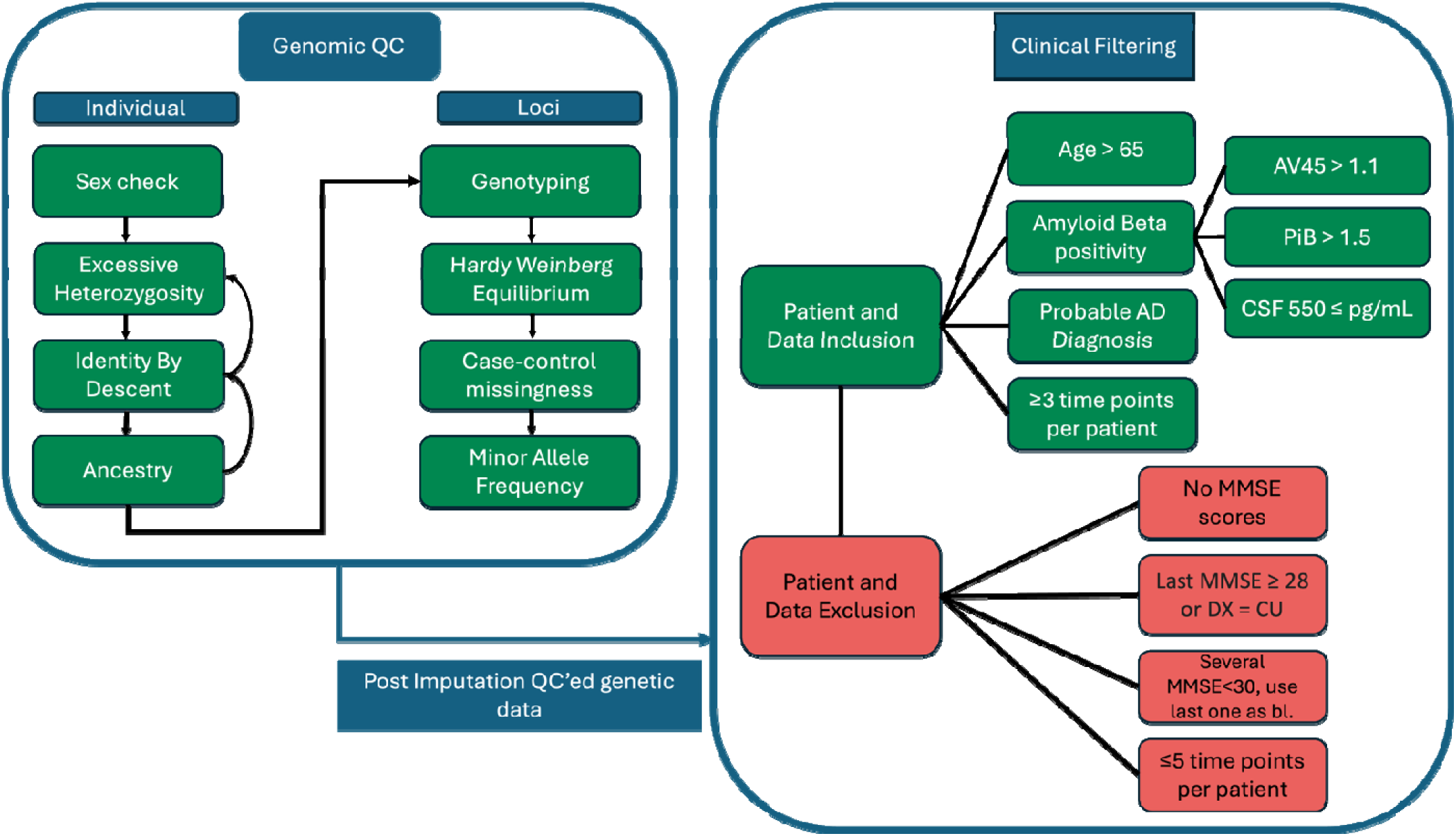
Filtering steps for genetic and clinical longitudinal data. Genomic QC was performed per individual first. Excessive heterozygosity checks, Identity by descent and ancestry checks were performed multiple times if needed. Clinical data filtering com rised of two components, Individual and data-based filtering with each comprising inclusion and exclusion criteria as shown. DX, bl and CU denote diagnosis, baseline, and cognitively unimpaired respectively.

### 2.3. Patient filtering

To model AD clinical progression, the ADNI and AIBL clinical data were filtered to retain only patients with probable AD and only data points capturing active decline, *i.e*. disease progression (Figure 1). To prevent inclusion of other dementias as AD, only Aβ-positive patients (likely to present amyloid plaque) were retained. For ADNI, amyloid status was assessed via neuroimaging data using ligands Pittsburgh compound B (PiB), and florbetapir F18 (AV45). In the absence of neuroimaging data, CSF biomarkers for Aβ were used. Here, patients with AV45 > 1.1, PiB > 1.5 or CSF Aβ ≤ 550 pg/mL were considered as Aβ-positive.

PET-amyloid imaging in the AIBL study uses five different tracers: AV45, PiB, ^18^F-flutemetamol (FMM), ^18^F-florbetaben (FBB), and ^18^F-NAV4694 (NAV4694 [25]). To align all values on the Centiloid (CL) scale, standardised uptake value ratios (SUVR) were calculated using the CapAIBL method [32] and these were transformed and expressed in CL using each tracer’s prescribed transform [33, 34]. A threshold of ≥ 21 CL was used to define amyloid positivity for this study [35].

Cognition was measured by MMSE score. Patients without MMSE scores, those with the most recent MMSE score ≥ 28 or those with the most recent diagnosis as “Cognitively unimpaired” (CU) were removed. For remaining patients, all data points before their most recent MMSE score of 30 were removed to ensure active decline. For patients with more than 5 visits, only the last (most recent) 5 were retained to avoid biassing mixed-effects models with wide variation in the number of data points per person. Finally, patients with < 3 data points were removed.

### 2.4. Mixed-effects modelling

A linear mixed-effects model with both random intercepts and slopes was built to capture within- and between-patient variation in MMSE scores over time. The model was chosen based on optimisation using on the ADNI clinical data by adding one variable at a time to maximise conditional R^2^ and minimise AIC.

The first visit per patient was set as baseline (Time = 0), then time in months was calculated from patient age at each visit. The highest conditional R^2^ (0.941) and lowest AIC on the ADNI data was returned from the following model, which was used for the polygeneic risk score, *APOE-*ε*4* and GWAS models.

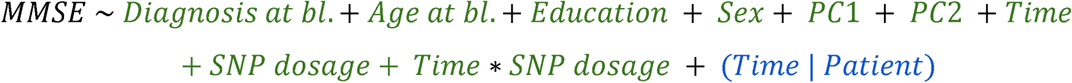

In this model the interaction term determines SNP effect on progression where SNP denotes any genetic component being analysed.

### 2.5 Power Calculations

To estimate the statistical power to detect an effect of the candidate SNP on the rate of cognitive decline, we performed a simulation-based power analysis using the *simr* package in R. We first fitted a linear mixed-effects model to our longitudinal MMSE data from ADNI for a random SNP as stated above with the modification from a correlated (Time | Patient) to (1 | Patient) + (0 + Time | Patient). This uncorrelated model was adapted for power calculations to address convergence issues.

Parameters for the simulation, including fixed-effect coefficients and variance components for the random effects, were derived from this initial model fit. To define clinically meaningful effect sizes for the SNP dosage×Time interaction term, we modelled scenarios where the SNP would increase the baseline rate of MMSE decline (observed in the SNP dosage=0 group) by 50%, 100% (doubling), and 200% (tripling). Power was calculated for each scenario based on 500 simulations per effect size, with a significance threshold of α=0.05.

### 2.6. Polygenic risk scores

Polygenic risk scores (PRS) were used to assess whether established genetic risk factors for AD might drive disease progression in our sample. Methods previously described by Altmann et al. [36] were adapted in PRS calculation based on SNP effect sizes from a meta-analysis of AD risk by the Alzheimer’s Disease Genetics Consortium (ADGC; [37]). Importantly, this selection avoids the reduction in SNP-based heritability that is seen in more recent AD GWASs [38]. The *APOE* region was removed from the Kunkle et al. summary statistics [37] and LD-clumping (1MB, R^2^ of 0.1 and P-value threshold of 1.0) was performed in PLINK v1.9 [27]. PRSice v2.1.9 [39] was then used to generate PRSs with thresholds of *p*≤ 5 ×10^-8^, (PRS_GW_; nSNPs =8) *p*≤ 5 ×10^-5^ (PRS_Nominal_; nSNPs = 11,723), and *p*≤ 0.5 (PRS_0.5_; nSNPs = 57,055). These scores were adjusted by the *APOE-*ε*2* and *APOE-*ε*4* estimated effect sizes in the Kunkle et al. GWAS [37] (*i.e.*, -0.467 & 1.202, respectively). Finally, analyses were conducted on the pooled ADNI and AIBL dataset using an equivalent linear mixed-effects model as above (Section 2.4) but with the SNP dosage term replaced with each of the three PRSs.

### 2.7. GWAS

The linear mixed-effects model was run on every SNP in the ADNI data and on ADNI-overlapping SNPs in the AIBL data, individually then meta analysed using METAL, weighted by cohort sizes [40] using standard error and effect sizes.

### 2.8 Functional Annotation

#### 2.81 Primary Functional Annotation

Summary statistics were analysed through the FUMA *SNP2GENE* pipeline [41], using a significance threshold of *p*≤ 5 ×10^-5^ to identify suggestively significant results. The following steps were only performed for SNPs at *p*≤ 5 ×10^-5^: SNPs in LD with candidate SNPs were mapped using the 1000 Genomes Phase 3 European cohort as a reference panel [31]. Genes within 10kb of candidate and lead SNPs were mapped to infer their potential functional significance. Significant expression quantitative trait loci (eQTLs) were identified among candidate SNPs using GTEx/v8 bulk tissue expression data [42] at *p*≤ 5 ×10^-5^ for high confidence.

Finally significant 3D chromatin interactions were identified using PsychEncode, GSE87112, and hESC within 250-500 KB of the candidate SNPs, with false discovery rate filter of 1×10 ^-6^.

#### 2.8.2 Ingenuity Pathway Analysis

Ingenuity Pathway Analysis (IPA, Qiagen Inc.) was performed using genes mapped to lead SNPs (*p*≤ 5 ×10^-5^) through positional mapping, or functional annotations. The magnitude of the corresponding genome wide regression coefficients were used as the strength of association. The *Core Analysis* was run with default settings, using the Ingenuity Knowledge Base (human genes only) as background. Pathway enrichment was assessed using activation z-scores and one-sided Fisher’s exact test to evaluate the overrepresentation of GWAS-prioritised genes in canonical pathways relative to the background gene set.

#### 2.8.3 Cell Type Enrichment Analysis

GWAS-prioritized progression genes were tested for enrichment across brain cell types using a one-sided Fisher’s exact test (hypergeometric test) and optimised using guidelines from Botía *et al.* [43]. The background gene set was defined as all human protein-coding genes (∼20,000). Cell type specific gene sets were obtained from the *CoExpNets* R package [44], which compiles markers from published single-cell datasets [43, 45]. P-values were adjusted for multiple testing using the Benjamini-Hochberg method, and cell types with a false discovery rate (FDR) below 0.05 were considered significantly enriched. Results were visualized as a heatmap of significant cell types.

### 2.9 Fine-mapping

Fine-mapping of each independent locus highlighted by FUMA, was analysed through statistical and functional fine-mapping of GWAS Risk loci mapping (*SAFFARI [46]*) which is designed to implement Snakemake and run four different fine-mapping methods (*SuSiE [47], FINEMAP [48], PolyFun+SuSiE, PolyFun+FINEMAP*). UK Biobank linkage disequilibrium matrices were used due to the larger sample sizes. Briefly, *PolyFun* [49] is a computational framework that improves the accuracy of fine-mapping by incorporating functional annotations across the entire genome to specify prior probabilities for fine-mapping methods like *SuSiE* and *FINEMAP*. It surpasses previous fine-mapping methods by using genome-wide data, estimating functional enrichment with a broad annotation set, and specifying prior causal probabilities based on predicted per-SNP heritability.

### 2.10 Co-localisation Analysis

Co-localisation was performed using *coloc* in R, on each independent locus (*p*≤ 5 ×10^−5^) highlighted by FUMA against MetaBrain eQTLs from five brain regions: Cortex, Hippocampus, Basal Ganglia, Spinal cord and the Cerebellum. The cis-eQTLs were obtained from https://metabrain.nl/cis-eqtls.html for the European cohort and processed per locus.

## 3. Results

### 3.1. Clinical data distribution of cohort

A total of 222 ADNI and 165 AIBL participants were included in the final sample after patient filtering. Their data was pooled for the 3,560,410 SNPs that were identified as overlapping across both QCed cohort-specific datasets. Distributions of clinical variables for those in the final dataset were then reviewed and differences across cohorts were identified (Table 1). On average, ADNI participants were more likely to be male, to have a greater number of years of education, to be younger at baseline, and to show a faster decline in MMSE than AIBL participants. While ADNI participants had a slightly higher average number of visits than AIBL participants, those in AIBL had a considerably longer average participation duration.

**Table 1:** Clinical Characteristics by Study Cohort

| Variable | Levels | ADNI | AIBL | p |
| --- | --- | --- | --- | --- |
| Sex | Female | 90 (40.5) | 85 (51.5) | 0.041* |
|  | Male | 132 (59.5) | 80 (48.5) |  |
| Education (Years) | Mean (SD) | 15.7 (2.8) | 12.1 (2.5) | <0.001*** |
| Age at Baseline (Years) | Mean (SD) | 72.9 (7.4) | 74.9 (6.7) | 0.008* |
| MMSE at Baseline | Mean (SD) | 25.4 (2.8) | 24.9 (4.1) | 0.184 |
| Number of Visits | Mean (SD) | 4.3 (0.7) | 4.0 (0.9) | <0.001*** |
| Duration of Participation (Months) | Mean (SD) | 35.4 (16.6) | 57.5 (17.3) | <0.001*** |
| Change in MMSE per year | Mean (SD) | -2.2 (2.3) | -1.7 (1.5) | 0.022 |
| APOE ε4 Dosage (n) | 0 | 59 (26.6) | 52 (31.5) | 0.36 |
|  | 1 | 114 (51.4) | 85 (51.5) |  |
|  | 2 | 49 (22.1) | 28 (17.0) |  |
| Diagnosis at Baseline (n %) | CN | 9 (4.1) | 46 (27.9) | <0.001*** |
|  | MCI | 119 (53.6) | 56 (33.9) |  |
|  | AD-Dementia | 94 (42.3) | 63 (38.2) |  |

This likely reflects the fact that AIBL participants are scheduled for visits at 18-month intervals [24, 25] whereas ADNI clinical visits are typically spaced by 6 to 12 months [22]. More than half of those from ADNI (53.6 %) were classified as MCI at baseline and few (4.1%) were CU, with the remaining 42% being AD-dementia. This was significantly different to AIBL participants where baseline classifications were distributed evenly between CU, MCI and dementia. No differences were observed in *APOE*–ε*4* allele carriage or baseline MMSE scores across cohorts.

MMSE values were right skewed towards the test ceiling of 30 for both studies, although a greater spread in the distribution was seen for AIBL (Figure 2A). Average scores at baseline show reductions with advancing diagnostic categories, as expected (Figure 2B). While these appear reasonably consistent across studies, those with a baseline classification of dementia in the AIBL study had notably more variation in corresponding MMSE scores than those with the same baseline classification in ADNI.

**Figure 2:**
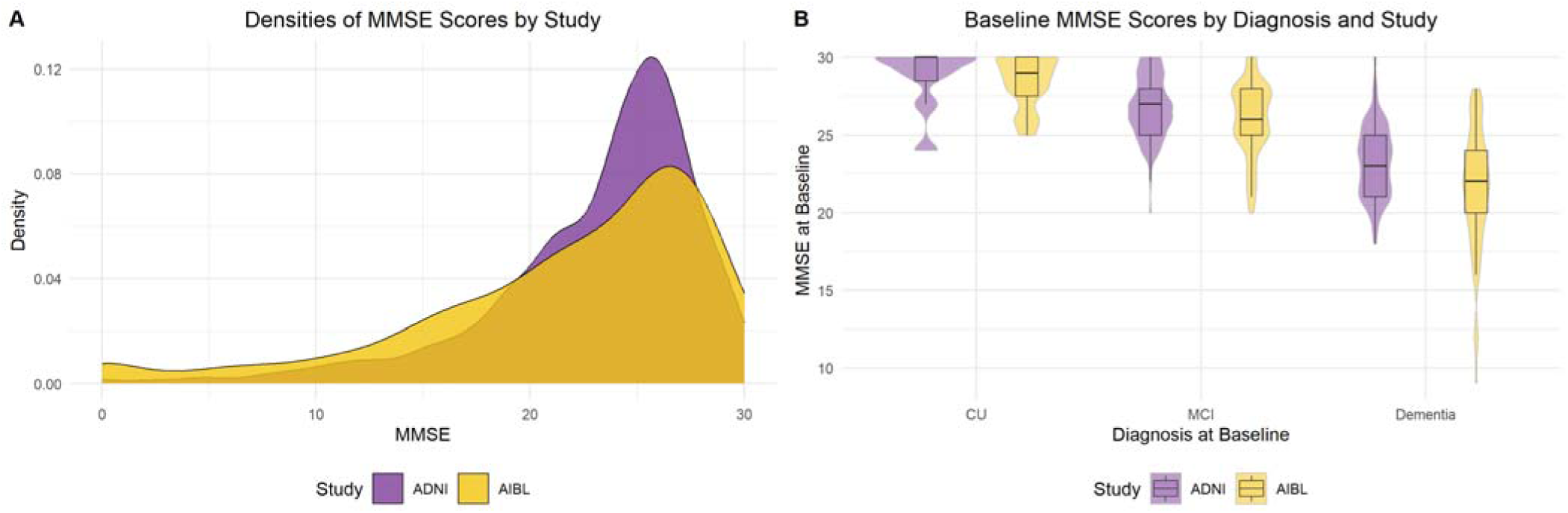
Clinical data distribution of ADNI and AIBL cohorts. (A) Distribution of MMSE scores across the longitudinal datasets, (B) MMSE at baseline by diagnosis at baseline (where CU = cognitively unimpaired and MCI = mild cognitive impairment).

### 3.2 Power Analysis

The power analysis performed on ADNI samples alone, revealed that the study was well-powered to detect variants conferring a severe effect on the rate of cognitive decline but was underpowered for smaller effects.

Previous studies using the PRS models used here, indicated an odds ratio (OR) of 1.9 for AD risk[50], whilst the effect of *APOE-*ε*4* on AD risk within Caucasian populations has been quoted as an OR of approximately 3.0.

The ADNI samples alone provided a 28.4% power to detect a variant that increased the rate of decline by 50% (interaction β=−0.068 MMSE points/month). The power to detect a doubling of the decline rate on par with the effect of polygenic risk scores on AD risk, (interaction β=−0.136 points/month) was 74.4%. For a variant that tripled the rate of cognitive decline (interaction β=−0.272 points/month), a magnitude analogous to the threefold increase in disease risk conferred by *APOE-*ε*4* in Caucasian populations, the power calculations indicated 100% power (95% CI: 99.3-100%). These results suggest that the longitudinal design used here had robust statistical power to identify effects of PRS and *APOE*–ε*4* in our data, if they were to have similar effects in progression as risk. It also implies that should genetic variants with moderate effects on progression exist, they would be identifiable in our combined cohort of ADNI and AIBL.

### 3.3. APOE

Results from the mixed-effects model testing effect of *APOE*–ε*4* dosage on rate of change in MMSE in the pooled sample are shown in Table 2. No lone effect (β=-0.046, p=0.811) for *APOE*–ε*4* or interaction with months since baseline (β=-0.016, p=0.143) was seen in the data. Given the main effect of *APOE*–ε*4* on clinical disease is through its influence on amyloid clearance [51, 52]. the absence of a role for *APOE*–ε*4* on progression is concordant with the inclusion criteria of amyloid positivity at baseline.

**Table 2:** Mixed effects modelling of APOE-ε4 on AD progression in pooled dataset

|  | Estimate | Std. Error | z-value | Pr(>\|z\|) |
| --- | --- | --- | --- | --- |
| Baseline Diagnosis (Dementia) | -6.159 | 0.419 | -14.964 | <0.001*** |
| Baseline Diagnosis (MCI) | -2.162 | 0.407 | -5.314 | <0.001*** |
| Age at Baseline | -0.021 | 0.019 | -1.109 | 0.268 |
| *APOE-ε4* Dosage | -0.046 | 0.191 | -0.239 | 0.811 |
| Education (Years) | 0.089 | 0.043 | 2.065 | 0.039* |
| Sex (Male) | 0.429 | 0.269 | 1.592 | 0.111 |
| PC1 | -1.448 | 2.562 | -0.565 | 0.572 |
| PC2 | -0.966 | 2.527 | -0.382 | 0.702 |
| Months Since Baseline | -0.016 | 0.011 | -1.466 | 0.143 |

This analysis also confirmed some covariate effects at baseline on the average MMSE. Specifically, poorer performance was seen in those with a diagnosis of dementia (β=116.159, p<0.001) or MCI (β=-2.162, p<0.001). Higher education was associated with higher average change in MMSE (β=0.089, p=0.039). No significant effects were seen for age at baseline, sex, or the top two genetic principal components.

### 3.4 Polygenic risk scores

In the polygenic model, the variance explained by the various PRS as measured by R^2^ [53] did not change significantly, and Bayesian Information Criterion (BIC) differed only marginally between the models (Table 3), favouring the smaller set of SNPs.

**Table 3:** Mixed effects modelling of AD polygenic risk score effects on AD progression. Model name represents the p-value threshold from Kunkle et al summary statistic. Beta (β) is the natural log of the coefficient.

| Name |  |  |  |  |  |
| --- | --- | --- | --- | --- | --- |
| p-value threshold (nSNPs) | BIC | Parameter | Beta | Conditional-Marginal R2 | P-value |
| Genome-wide |  |  |  |  |  |
| $p \leq 5 \times 10^{-8}$ | 8176 | PRS*time(months) | -0.009 | 0.882 – 0.380 | .0294 |
| -8 |  | PRS | 0.126 |  | 0.412 |
| Nominal |  |  |  |  |  |
| $p \leq 5 \times 10^{-5}$ | 8182 | PRS*time(months) | -0.001 | 0.882 – 0.377 | 0.554 |
| -10598 |  | PRS | -0.023 |  | 0.56 |
| Legacy |  |  |  |  |  |
| $p \leq 0.5$ | 8182 | PRS*time(months) | -0.002 | 0.882 – 0.377 | 0.132 |
| -51964 |  | PRS | -0.017 |  | 0.479 |

### 3.5 Meta-analysis

The meta-analysis (Figure 3A and 3B) found one genome-wide significant locus (p ≤ 5×10*^-8^*) on chromosome 22 (rs78369883, p=2.06×10^-8^, β=0.191) and 25 nominally significant hits (p ≤ 5*×10^-5^*) detailed in Table 4.

**Figure 3.**
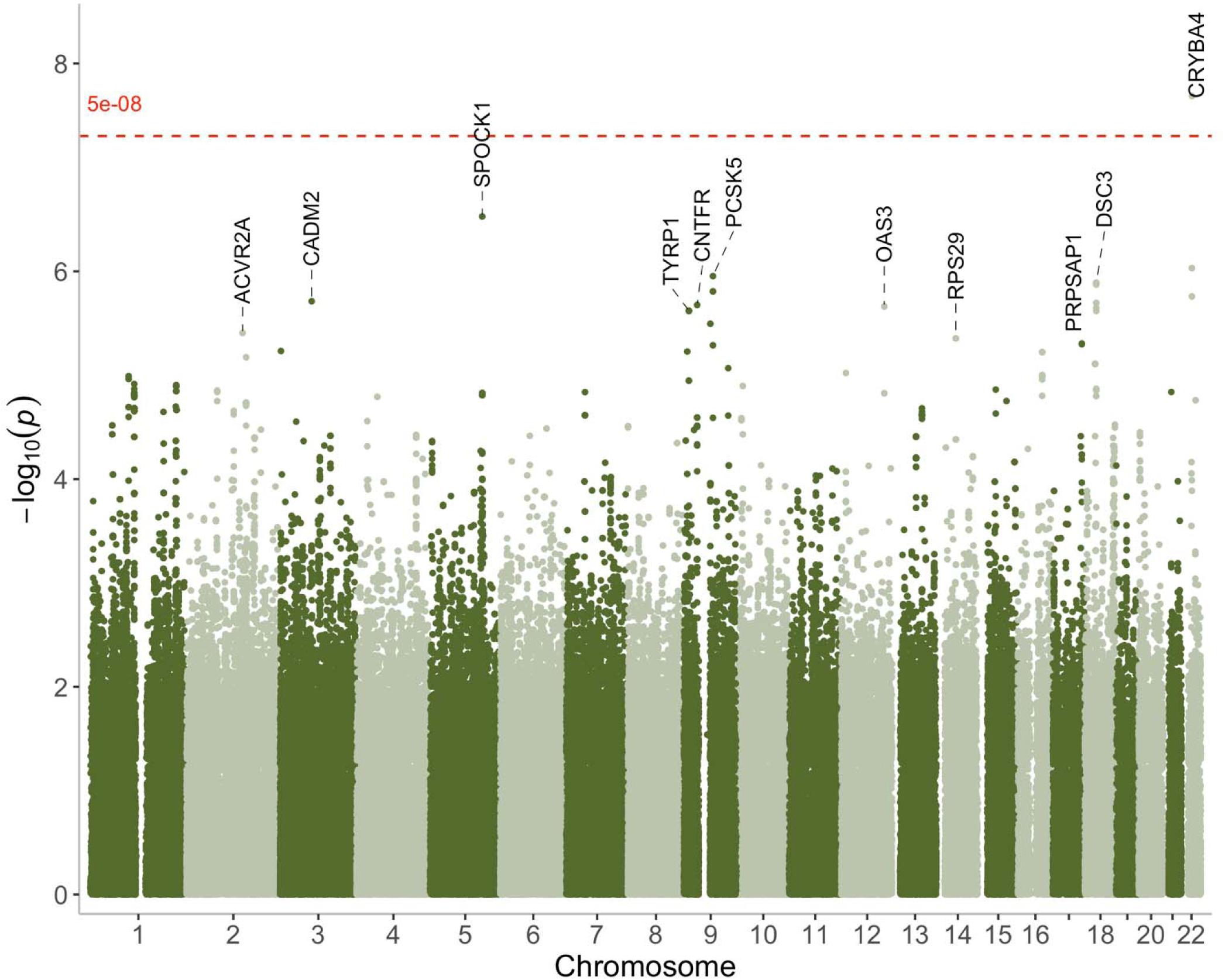
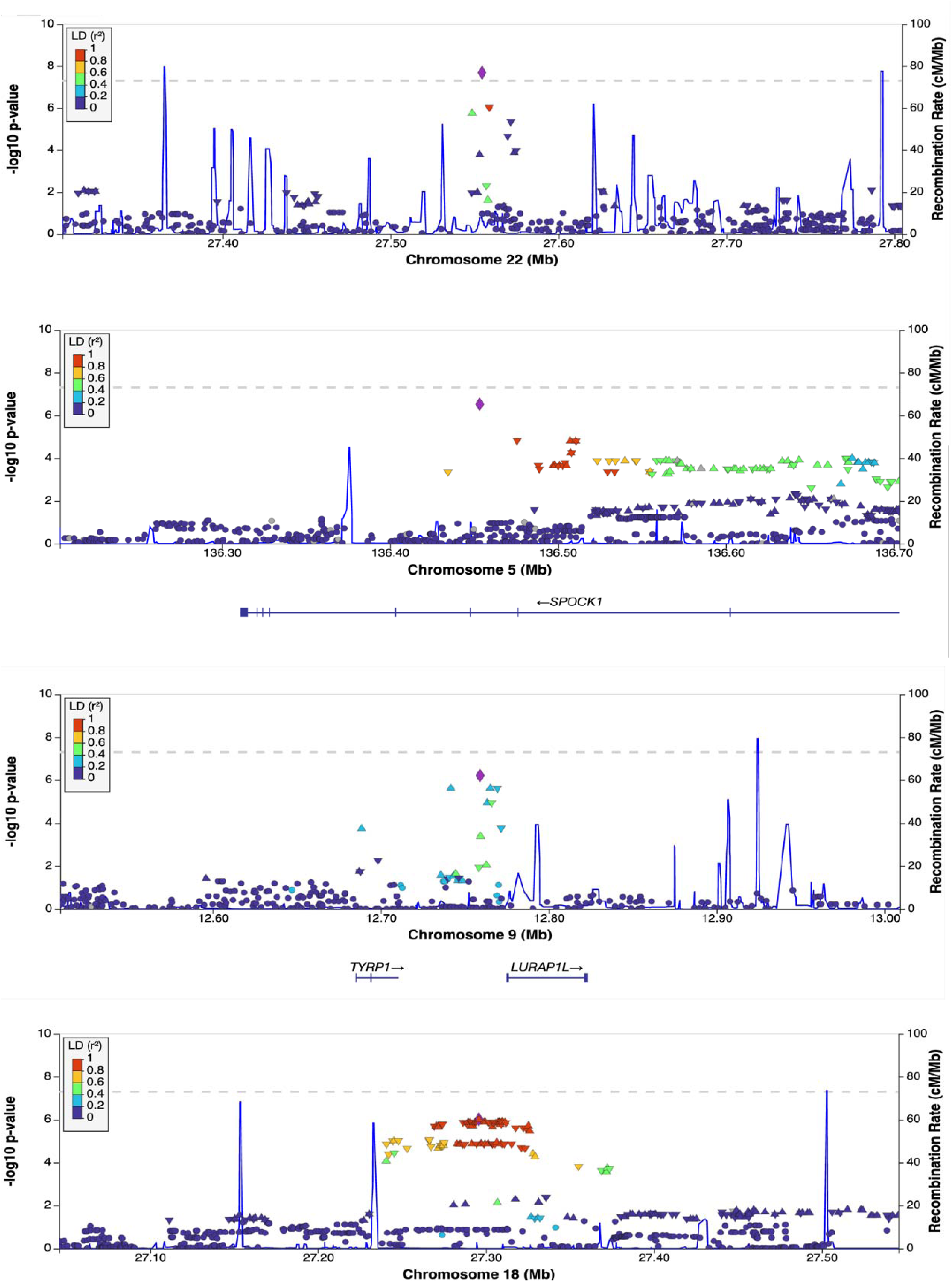
a) Manhattan plot of meta-analysis and pooled GWAS. b) locus zoom plots for top four loci.

**Table 4:** Summary statistics of meta-analysis independent signals at p<5×10-5. CHR is chromosome, BP is position, EA is the effect allele for which beta and p-value are shown. Beta and P-value denote the coefficient and p-value of the SNP*time parameter. Position coordinates are given in GRCh37.

| CHR:BP | SNP | Effect Allele (EA) | Non-EA | EA freq | Beta | P-value | Implicated genes |
| --- | --- | --- | --- | --- | --- | --- | --- |
| 22:27554525 | rs78369883 | T | C | 0.025 | 0.191 | $2.06 \times 10^{-8}$ | *MN1*,<br>*PITPNB*,<br>*ZNRF3* |
| 5:136453478 | rs61511032 | T | C | 0.025 | 0.178 | $2.97 \times 10^{-7}$ | *TRPC7*,<br>*SPOCK1*,<br>*SMAD5*,<br>*SLC25A48*,<br>*IL9* |
| 9:12759133 | rs60081651 | A | G | 0.04 | 0.134 | $6.08 \times 10^{-7}$ | *TYRP1*,<br>*LURAP1L* |
| 18:27295720 | - | A | ATC | 0.35 | -0.053 | $9.51 \times 10^{-7}$ | *CDH2*,<br>*DSC3* |
| 9:78792181 | rs111866233 | T | G | 0.013 | 0.227 | $1.11 \times 10^{-6}$ | *PCSK5* |
| 9:34548138 | rs77364997 | T | C | 0.04 | 0.131 | $2.11 \times 10^{-6}$ | *IL11RA*,<br>*GALT*,<br>*CNTFR* |
| 12:113409278 | rs45575939 | T | C | 0.024 | 0.162 | $2.18 \times 10^{-6}$ | *HECTD4*,<br>*RPL6*,<br>*OAS1*,<br>*OAS3* |
| 9:71884233 | rs73452954 | A | G | 0.118 | -0.078 | $2.35 \times 10^{-6}$ | *PIP5K1B*,<br>*TJP2*,<br>*C9orf135* |
| 2:148304479 | rs116515081 | A | G | 0.016 | 0.194 | $3.92 \times 10^{-6}$ | *ACVR2A*,<br>*ORC4*,<br>*LYPD6B*,<br>*LYPD6* |
| 14:49563156 | rs7144266 | T | G | 0.039 | 0.124 | $4.42 \times 10^{-6}$ | *RPL10L*,<br>*MDGA2*,<br>*RPL36AL* |
| 16:63787701 | rs112062540 | C | G | 0.113 | 0.077 | $4.98 \times 10^{-6}$ | *CDH11* |
| 3:2470968 | rs150742532 | T | G | 0.9 | -0.078 | $5.85 \times 10^{-6}$ | *LSAMP*,<br>*CNTN4*,<br>*IL5RA* |
| 11:79762744 | rs7119783 | A | G | 0.377 | 0.046 | $6.32 \times 10^{-6}$ | *TENM4* |
| 2:157782408 | rs10185072 | T | C | 0.067 | 0.095 | $6.71 \times 10^{-6}$ | *NR4A2*,<br>*GALNT5* |
| 16:53779005 | rs2388256 | A | G | 0.217 | 0.057 | $9.47 \times 10^{-6}$ | *RBL2*,<br>*RPGRIP1L*,<br>*AKTIP*,<br>*FTO* |
| 1:116417733 | rs1418662 | A | G | 0.962 | -0.117 | $1.21 \times 10^{-5}$ | *SLC22A15*,<br>*MAB21L3*,<br>*NHLH2*,<br>*TSPAN2* |
| **1:226824217** | rs1151804 | T | C | 0.114 | 0.068 | 1.24×10-05 | *ADCK3, ITPKB, C1orf95, PARP1, PSEN2* |
| **8:252095** | rs12547882 | A | G | 0.014 | 0.194 | 1.55×10-05 | - |
| **22:37183484** | rs17748070 | T | C | 0.021 | 0.17 | 1.74×10-05 | *IFT27* |
| **3:116352025** | rs61419876 | T | G | 0.055 | 0.097 | 2.06×10-05 | *LSAMP, UPK1B* |
| **1:227586014** | rs146787643 | T | G | 0.985 | -0.195 | 2.16×10-05 | *ADCK3, JMJD4, ITPKB, C1orf95, SNAP47, PSEN2, H3F3A, CDC42BP A* |
| **6:78494039** | rs117110256 | A | C | 0.017 | 0.171 | 3.83×10-05 | *IRAK1BP1, HMGN3, HTR1B, MEI4* |
| **22:26417956** | rs761670 | T | C | 0.455 | 0.043 | 6.85×10-05 | *MYO18B, SEZ6L, TFIP11* |

### 3.6 Implicated loci

The 26 loci map to thirteen chromosomes, with notable importance of hits on chromosomes 22, 18, 12, and 5.

The loci identified within these signals mapped to gene-rich regions. Expression quantitative trait loci, splicing quantitative trait loci, and chromatin loop interactions, provide evidence of a role for the implicated genes. A total of 180 genes were identified in these regions, prompting fine-mapping techniques and colocalization analysis to further refine the identified loci.

The lead SNP on chromosome 22 (rs78369883, CADD= 14), located in intron 1 of long non-coding RNA CTA-992D9.7 reached genome-wide significance. It is predicted to be deleterious by CADD and is in LD (D’ ≥ 0.8) with SNPs notably associated with longitudinal cognition [54], high-sensitivity cardiac troponin I concentration [55] visual disorders [56, 57] and mean bone mineral density[58]. The SNP is at or near an Active Transcription Start Site on the chromatin state in the Anterior Caudate and is likely to affect the transcription of CTA-992D9.7. CTA-992D9.7 (ENSG00000231405) is expressed primarily in the brain with the highest levels of expression in the caudate, higher than any other human tissue (ARCHS4)

The chromosome 18 hit at position 27295720 (rs74546314) predicted to be deleterious (CADD = 17), is linked by chromatin loop interactions to *CDH2* and *DSC3*. *CDH2* encodes N-cadherin, a cell adhesion molecule crucial for neuronal connections and synaptic plasticity and has been implicated in neurodevelopmental pathways involving Reelin signalling, and also neurodegenerative diseases [59]. SNPs in high LD (D’=0.9) with this SNP have shown association with traits such as Neurofilament light chain levels, MMSE scores, Global Cognition, Apolipoprotein A1 levels, and Intelligence.

Rs45575939 on chromosome 12 notably maps to *OAS3*. The OAS family (*OAS1*, *OAS2 and OAS3*) form part of the same haplotype directly associated with AD and involved in the interferon response linked with neuroinflammation and synapse loss in AD [60] and linked with a pro-inflammatory response via STAT/NF-kappaB in microglia/macrophages when experimentally downregulated [61]. This is the only haplotype that has been associated with both risk and progression.

*SPOCK-1* was implicated through chromatin interactions on chromosome 5 (5:136453478, rs61511032 CADD=19) and encodes Testican-1, which is expressed by neurons and oligodendrocytes. It is involved in cell-cell and cell-matrix interactions and plays a role in the development and function of the blood-brain barrier [62]. It is known to activate NF-kappaB, Wnt/β-catenin, PI3K/Akt, and mTOR/S6K pathways and as such can heavily influence neurodegenerative mechanisms.

Fine-mapping analysis using *SAFFARI*, identified SNP rs116515081 on chromosome 2, mapped to *ACVR2A* (2:148304479) as a likely causal contributor to progression in AD, with a posterior probability greater than 80% by both *PolyFun* and *FINEMAP*. *FINEMAP* provides purely statistical evidence for this association, while *PolyFun* leverages functional genomic annotations to enhance the statistical model. *ACVR2A* plays a crucial role in oligodendrocyte differentiation and axonal ensheathment [63]

Colocalisation analyses indicate that the locus at 5:136453478 demonstrates significant colocalisation with eQTLs for *SLC25A48* and *IL9* in the cortex, evidenced by posterior probabilities (PP.H4) of 70.0% and 62.0%, respectively. Similarly, loci 1:227586014 and 1:226824217 exhibit colocalization with the eQTL for *MIXL1* (also known as *MILD1*) within the cortex, each with a posterior probability of colocalization of 62 and 63%. *MIXL1*, is a homeobox transcription factor which plays a role in embryonic development in mesol1J and endoderm stages.

### 3.6 Implicated Pathways

Analysis using *gprofiler2* identified the genes at these 26 loci as significantly enriched in a variety of biological annotations and Gene Ontology terms including cellular zinc and copper responses, oligoadenylate synthetase activity, neuron projection, cell membrane projection, cellular homeostasis, interleukin-5 activity, cytokine-receptor interactions, and obesity (Figure 4).

**Figure 4:**
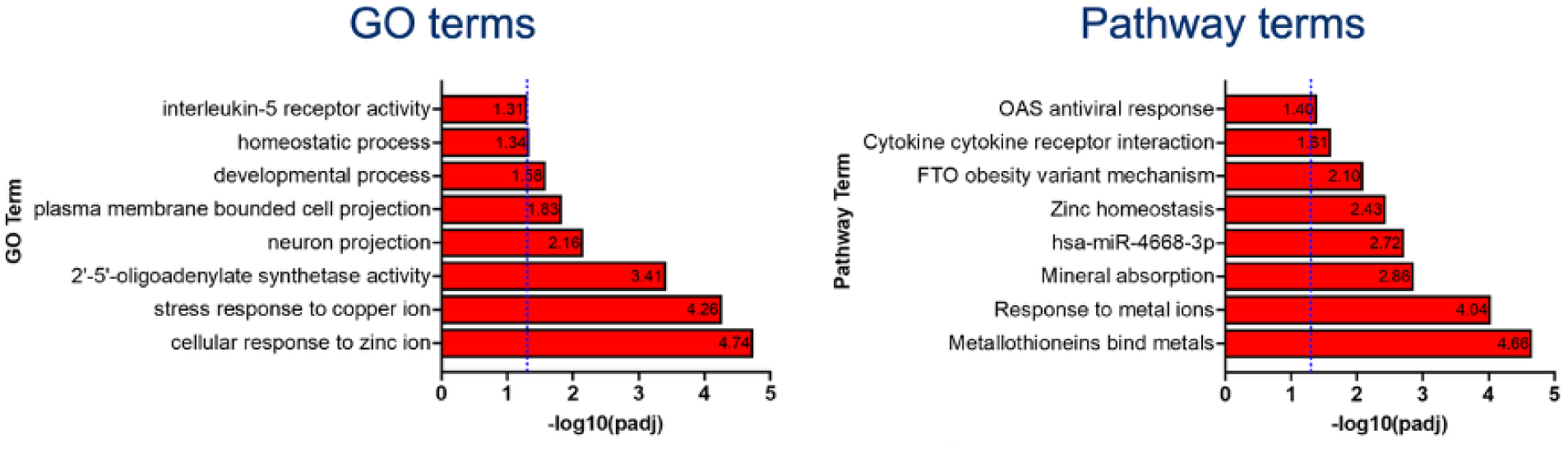
Enrichment of AD Progression loci in immune and neuronal processes: Annotation with gprofiler2.

Using IPA, we observe that these genes were enriched in pathways representing anti-viral, interferon, cancer, serotonin receptor, STAT3, NGF, BMP, circadian, interleukin-6 and senescence-related processes (IPA analysis; Figure 5). This suggests that these loci are involved in controlling a variety of pathways mediating neuronal-interferon-microglial signalling relating to normal homeostatic and housekeeping processes of neuronal networks and their development.

**Figure 5:**
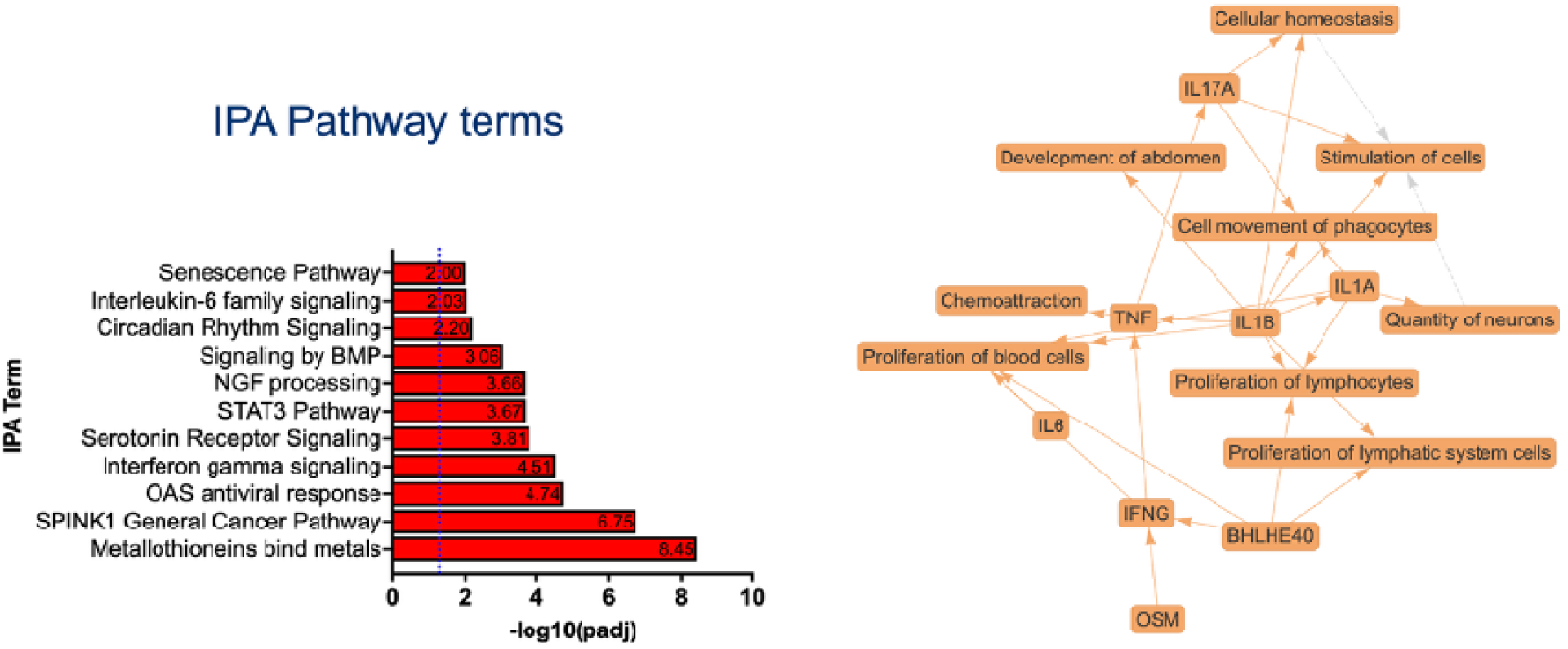
IPA analysis reveals enrichment of AD progression loci in immune and neuronal pathways. The graphical summary illustrates key pathways, regulator activation and biological function predictions from IPA Core Analysis, which enriched mainly in inflammatory pathways, and neuroimmune interactions.

Brain cell-type analysis revealed significant enrichment of progression genes in specific neuronal populations, notably dopaminergic and inhibitory subtypes, in both datasets studied (Figure 6). Despite dementia impacting various neuronal systems, dopaminergic pathways linked to reward, and attention seem particularly influential in this analysis of AD progression, potentially shortening survival time.

**Figure 6:**
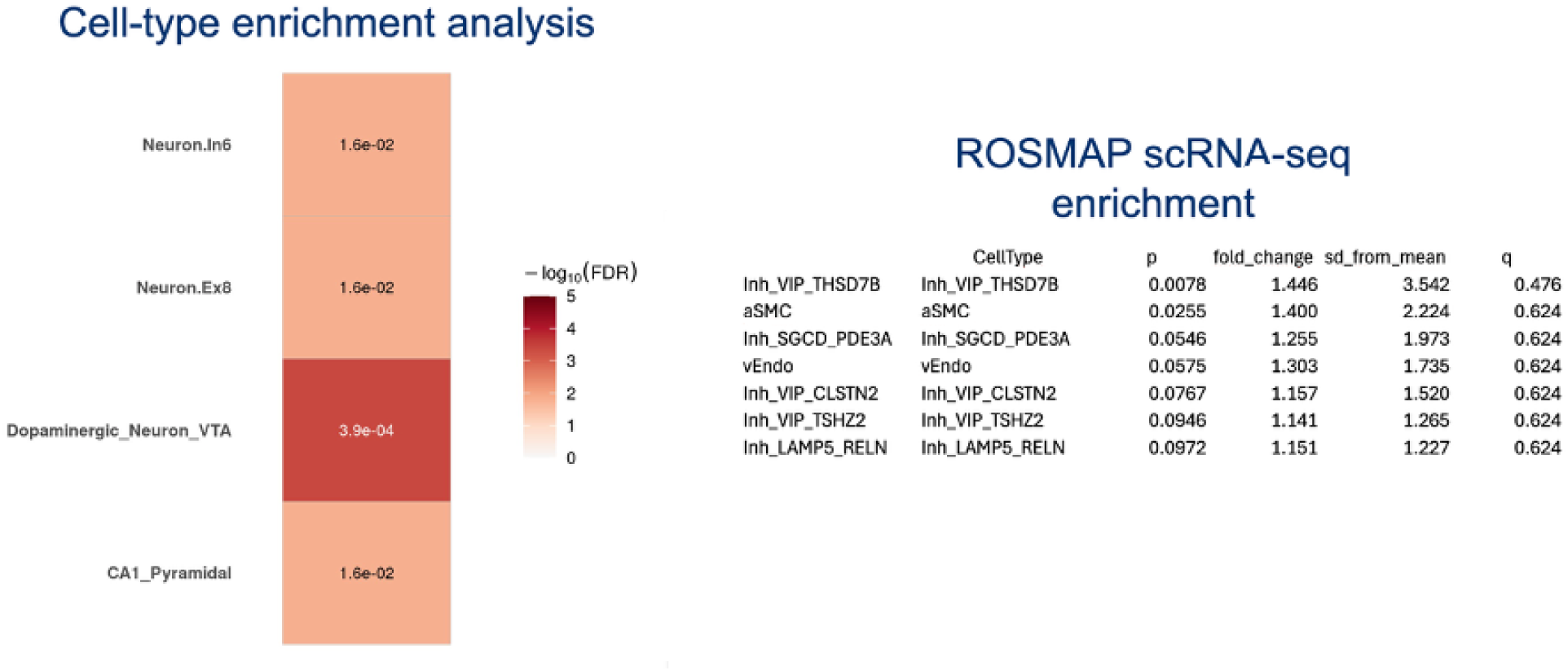
Cell-type enrichment analysis of AD progression loci reveals significant overrepresentation of differentially expresse genes in VTA dopaminergic and CA1 pyramidal neurons [43] and in VIP+ interneurons and aSMCs {Xiong, 2023 #516}{Skene, 2016 #517}

Analysis of loci that reach GWAS and nominal significance and SNPs in LD with them at D’ ≥ 0.8 reveals predominant associations with traits such as cognitive decline [17, 64, 65], educational attainment [66–69], mean bone density [58, 70], intelligence and performance intelligence quotient [71, 72] and soluble TREM-2 levels [73].

In summary, putative genes at loci associated with progression of AD are involved in pathways relating to tau accumulation, neuronal function and brain resilience, and immune response and inflammation as summarised in Table 5.

**Table 5:** Summary of pathways and upstream regulators implicated or predicted in progression of AD

| Pathway | Genes |
| --- | --- |
| Immune Response | *OAS1, NGF, CCL19, ANXA1, IL1RN, IFNG, IL9, BHLHE40, IL1A, IL1B, IL17A, IL6, TNF* |
| Tau/Neurodegeneration | *SPOCK-1, ZNRF3, CDH2, MMP2, PVALB, MAPK1, CASP6, PARP1* |
| Brain Resilience/<br>Neurogenesis | *CACNG2, CDH2, PITPNB, IRX3, IRX5, IRX6, SMAD5, TGFBI* |

## 4. Discussion

This study aimed to uncover genetic pathways that specifically influence Alzheimer’s disease progression rather than overall disease risk, and to identify potential targets to mitigate cognitive decline.

In these analyses we face three layers of uncertainty in the interpretation of the results. These are; the importance of SNPs that are approaching statistical significance, the implication of several genes at each loci, and the uncertainty in annotations available in the public domain. Nevertheless, several conclusions can be drawn with confidence. These are discussed below.

### 4.1. Uncoupling genetic factors of AD risk and progression

The primary role of the *APOE*–ε*4* allele in AD is to promote amyloid-β deposition [51, 52] through impairing its clearance. Since our cohort was exclusively amyloid positive, participants had already passed the pathogenic stage where *APOE*–ε*4* exerts its main effect and thus the lack of *APOE*–ε*4* dosage association with AD progression. We contend discrepancies seen in previous studies are explained by a common methodological limitation: the failure to use amyloid status to confirm AD diagnosis. This likely led to the confounding inclusion of non-AD patients, measuring the effect of *APOE*–ε*4* on disease risk rather than progression. Similarly, AD polygenic risk scores, which typically have been shown to be good predictors or AD risk, show a lack of association with progression, which further supports the notion that the genetic drivers of AD pathogenesis are distinct from those modulating progression.

### 4.2. Meta-analysis results

We performed a meta-analysis of AD progression on individual GWASs of a combined cohort of 387 adults with AD from the ADNI and AIBL studies to investigate potential drivers of AD related clinical disease progression and this identified one genome-wide significant locus (*p* ≤5 ×10^-8^) and 25 suggestively significant loci (*p* ≤5 ×10^-5^).

#### Associated Pleiotropic Traits of AD Progression Loci

Several common themes emerged in the analysis of SNPs in LD with our identified loci. Notably, cognitive decline and AD, immune response, ophthalmic disorders, and mean bone mineral density were implicated several times across different loci.

As an example, variants in high LD with rs78369883 (chromosome 22) and rs112062540 (chromosome 16) are associated with cognitive decline and altered delayed memory function in AD [54, 65], whilst rs17748070 (chromosome 22) and rs1151804 (chromosome 1) are linked to AD and sTREM-2 levels [73–75]. Eleven loci were linked to bone mineral density (BMD) [58].

#### Biological Functions of Implicated Genes

Multiple genes implicated in this study converge on pathways related to inflammation, tau, and the immune response.

Neuroinflammation emerges as a central theme, with several genes implicated in promoting inflammatory processes. This includes *ITPKB* which is associated with proinflammatory interleukin and microglial signalling in mouse models [76–79],*OAS3* which may regulate microglial function [80], and *TJP2* [81] which allows leakage of proinflammatory factors into the brain (and by extension highlights the role of Blood brain barrier dysfunction). Moreover, a potential shared innate immune pathway linking AD and BMD is known [70].

The findings illustrate a crucial interplay between neurodegeneration and neuroprotection. On one side, *ITPKB* and *TJP2* directly promote core neurodegeneration components, [76–79, 82, 83], while on the other, neuroprotective signalling pathways involving *CUTFR* and *IL11RA* reduce inflammation [84], support neurogenesis, and protect neurons [84, 85]. The receptor *CNTFR* binds CNTF, which can activate microglia [86], mitigate neurodegeneration in diseases including Huntington’s [87] and ALS [88], support myelin regeneration, protect oligodendrocytes, and counter their apoptosis [89–91]. IL-11 or CNTF signalling could thus be important for protecting the brain in neurodegenerative disease and may be dysregulated in fast AD progressors.

Finally, genes with broader regulatory roles were also identified. These include *ZNRF3*, which acts as a negative regulator of the Wnt signalling pathway, and *MN1*, which plays a role in transcriptional regulation during developmental processes.

### 4.4. Cell types implicated

The cell type analysis, with the caveat of the known limitations above, revealed that genes associated with the progression of AD are significantly enriched in specific neuronal populations. We therefore hypothesise that the decline following a diagnosis of AD is driven by distinct neuronal populations, pathways, and genes as seen in Figure 7. This key finding, validated across two independent datasets [43, 92], points specifically to the enrichment of these progression genes in dopaminergic and inhibitory neuron subtypes. Disease associated microglia (DAM) which were previously seen to be enhanced in AD pathogenesis [61, 80, 93] do not appear to play a substantial role in progression. Although AD is recognized for its widespread impact on neuronal systems, this analysis points to the vulnerability of dopaminergic pathways during decline, which are crucial for regulating attention and motivation.

**Figure 7:**
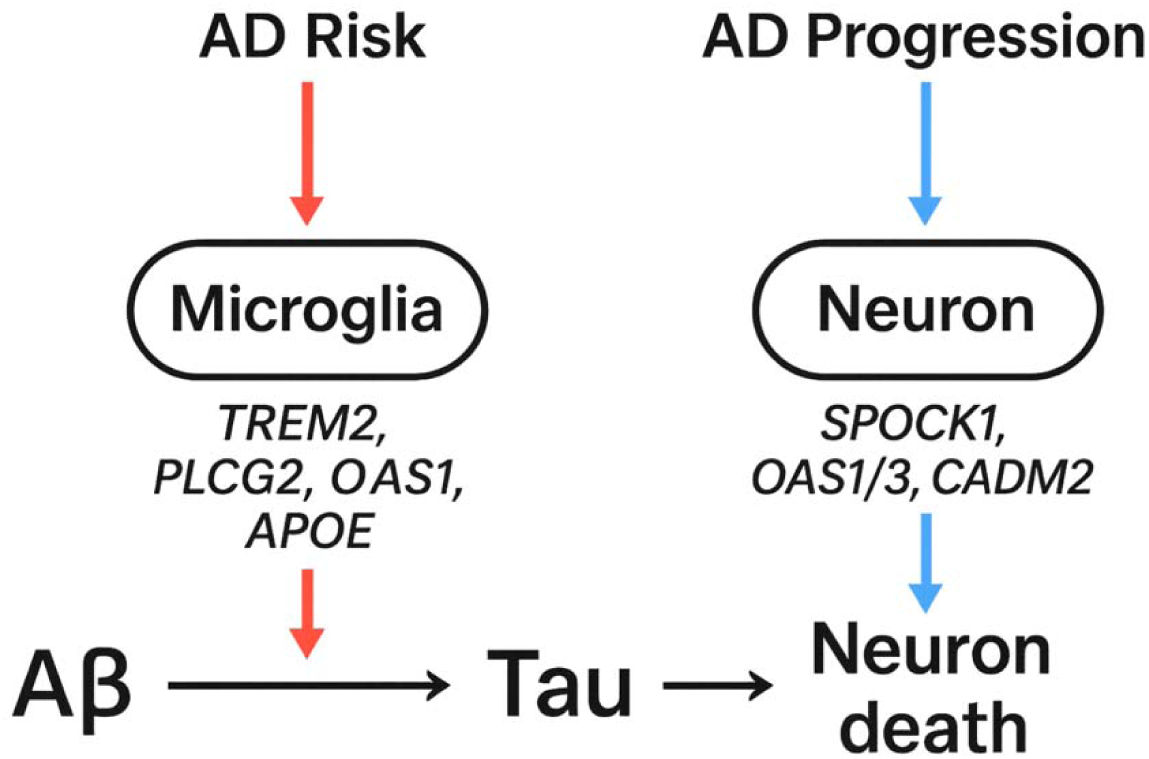
Proposed hypothesis of multistage involvement of neuronal cell types and pathways.

### 4.5. Study limitations and future direction

This study presented several limitations beyond those specified above, potentially impacting the significance and interpretation of results. Our data, in line with most publicly available cohorts, was biassed towards high MMSE values and low progression rates, potentially capturing the genetics of mainly early AD. While patient filtering was performed to limit this and retain only patient visits showing cognitive decline, more stringent filtering could be used in larger data (e.g., removing all patient visits with MMSE ≥28). Biases towards CU-like phenotypes could result in the mixed-effects model capturing AD risk rather than progression, but this is unlikely given that the main genetic AD risk factor *APOE*IZIε*4* has no effect.

Additionally, despite the stringent patient filtering and extensive phenotyping some patients may have other co-pathology, causing very fast, progression which cannot be excluded at this point. To mitigate this, future work with a larger dataset could remove the top 5% and bottom 5% of progressors after patient filtering as done in a Parkinson’s disease progression GWAS [94]. These extremes can be removed before mixed-effects modelling to avoid biassing the rest of the dataset.

Our small cohort whilst able to detect larger effect sizes is likely to miss smaller effects which may exert polygenic effects. Efforts are underway to undertake progression analyses in larger datasets from the placebo arm of clinical trials. The meticulous phenotyping and rigorous follow up data available from these trials will make these cohorts invaluable for progression studies. The larger dataset in these trials will also allow for inclusion of interaction with time for each of the covariates known to affect baseline status.

Finally, the interpretation of results could be influenced by confirmation bias in favour of known AD pathways (eg. amyloid aggregation, NFT formation, apoptosis, etc.) present in the existing literature and in our literature search, identifying existing links between mapped genes and AD.

To conclude, studying the genetics of AD progression has revealed much information about the biology of progression. This is likely activating through neuronal subtypes related to Tau pathology and neuronal resilience. Understanding these variants and pathways will provide targeted new opportunities for therapies around the time that MMSE scores start to decline and may provide new options for biomarker detection.

## Data Availability

The codes used for running the mixed effect models and the full resulting summary statistics are available upon request.

https://adni.loni.usc.edu/

https://aibl.org.au/

## Acknowledgements

We thank all the participants and their families who took part in both the AIBL and ADNI studies and the clinicians who may have referred participants. The AIBL study (www.AIBL.csiro.au) is a consortium between Austin Health, Commonwealth Scientific Industrial and Research Organisation (CSIRO), Edith Cowan University, the Florey Institute (The University of Melbourne), and the National Ageing Research Institute. A complete listing of AIBL investigators can be found at [https://aibl.org.au/about/our-researchers/].

The ADNI study was launched in 2003 as a public-private partnership led by Principal Investigator Michael W. Weiner, MD. For up-to-date information, see www.adni-info.org. We thank all the investigators within AIBL and the ADNI who may have contributed to the design, implementation, and coordination of these resources but did not actively participate in the development, analysis, interpretation or writing of this current study.

We wish to express our gratitude to Fidelity Bermuda Foundation, and Dolby Family Ventures for funding our work and their continuous support of our preliminary work.

## Funding

MS and JH received funding from Fidelity Bermuda Foundation, and Dolby Family Ventures for the work carried out in this manuscript.

Funding for this research was also supported through NHMRC grants (GNT1161706, GNT1191535) awarded to SML.

Data collection and sharing for AIBL is made possible through the support provided by study partners, including the CSIRO, Edith Cowan University (ECU), Mental Health Research Institute (MHRI), National Ageing Research Institute (NARI), Austin Health, and CogState Ltd.

The AIBL study has received funding from the National Health and Medical Research Council (NHMRC), the Dementia Collaborative Research Centres program (DCRC2), the Science and Industry Endowment Fund (SIEF), and the Cooperative Research Centre (CRC) for Mental Health – funded through the CRC Program, an Australian Government Initiative.

Data collection and sharing for ADNI is funded by the National Institute on Aging (National Institutes of Health Grant U19 AG024904). The grantee organization is the Northern California Institute for Research and Education. In the past, ADNI has also received funding from the National Institute of Biomedical Imaging and Bioengineering, the Canadian Institutes of Health Research, and private sector contributions through the Foundation for the National Institutes of Health (FNIH) including generous contributions from the following: AbbVie, Alzheimer’s Association; Alzheimer’s Drug Discovery Foundation; Araclon Biotech; BioClinica, Inc.; Biogen; Bristol-Myers Squibb Company; CereSpir, Inc.; Cogstate; Eisai Inc.; Elan Pharmaceuticals, Inc.; Eli Lilly and Company; EuroImmun; F. Hoffmann-La Roche Ltd and its affiliated company Genentech, Inc.; Fujirebio; GE Healthcare; IXICO Ltd.; Janssen Alzheimer Immunotherapy Research & Development, LLC.; Johnson & Johnson Pharmaceutical Research \& Development LLC.; Lumosity; Lundbeck; Merck & Co., Inc.; Meso Scale Diagnostics, LLC.; NeuroRx Research; Neurotrack Technologies; Novartis Pharmaceuticals Corporation; Pfizer Inc.; Piramal Imaging; Servier; Takeda Pharmaceutical Company; and Transition Therapeutics.

## Ethical Approval

The AIBL study is approved by ethics committees at St Vincent’s Health, Hollywood Private Hospital, Edith Cowan University, and The Commonwealth Scientific and Industrial Research Organisation (CSIRO). ADNI has ethics approval from all Institutional Review Boards where data was collected over the course 146 of the study. Ethics approval was also provided by Curtin University for the current analyses.

## Authors’ contributions

JH and MS: conceived the project.

SF, CEC, ARE, UY, and MS performed the analyses. ARE, MS, CEC developed scripts for analyses

TP, JH, SML, and DS: Supervised analysis. CEC and MS wrote the manuscript.

All authors contributed to manuscript editing.

## Data Access statement

The Alzheimer’s Disease Neuroimaging Initiative (ADNI) and the Australian Imaging, Biomarkers and Lifestyle (AIBL) study of aging are publicly available and can be accessed upon request. ADNI data are available via the LONI Image and Data Archive (IDA LONI). AIBL data are available via expression of interest to AIBL data access committee.

## Notes

### Competing Interest Statement

The authors have declared no competing interest.

### Author Declarations

The ADNI and AIBL studies are publicly available and can be accessed upon request. ADNI data are available via the LONI Image and Data Archive. AIBL data are available via expression of interest to AIBL data access committee. The AIBL study is approved by ethics committees at St Vincents Health, Hollywood Private Hospital, Edith Cowan University, and The Commonwealth Scientific and Industrial Research Organisation (CSIRO). ADNI has ethics approval from all Institutional Review Boards where data was collected over the course of the study. Ethics approval was also provided by Curtin University for the current analyses.

